# Impaired learning, memory, and extinction in posttraumatic stress disorder: translational meta-analysis of clinical and preclinical studies

**DOI:** 10.1101/2021.07.19.21260790

**Authors:** Milou S.C. Sep, Elbert Geuze, Marian Joëls

## Abstract

**Background:** Current evidence-based treatments for post-traumatic stress disorder (PTSD) are efficacious in only part of PTSD patients. Therefore, novel neurobiologically-informed approaches are urgently needed. Clinical and translational neuroscience point to altered learning and memory processes as key in (models of) PTSD psychopathology. We extended this notion by clarifying at a meta-level i) the role of information valence, i.e. neutral versus emotional/fearful, and ii) comparability between clinical and preclinical phenotypes. We hypothesized that, cross-species, neutral versus emotional/fearful information processing is, respectively, impaired and enhanced in PTSD.

**Methods:** This preregistered meta-analysis involved a literature search on PTSD+Learning/Memory+Behavior, performed in PubMed. First, the effect of information valence was estimated with a random-effects meta-regression. Then sources of variation were explored with a random forest-based analysis.

**Results:** The analyses included 92 clinical (N=6732 humans) and 182 preclinical (N=6834 animals) studies. A general impairment of learning, memory and extinction processes was observed in PTSD patients, regardless of information valence. Impaired neutral learning/memory and fear extinction were also present in animal models of PTSD. Yet, PTSD enhanced fear/trauma memory in preclinical studies and impaired emotional memory in patients. Clinical data on fear/trauma memory was limited. Mnemonic phase and valence explained most variation in rodents but not humans.

**Conclusions:** Impaired neutral learning/memory and fear extinction show very stable cross-species PTSD phenotypes. These could be targeted for novel PTSD treatments, building on neurobiological animal studies. We argue that seemingly cross-species discrepancies in emotional/fearful memory deserve further study; until then animal models targeting this phenotype should be applied with care.

## 1. Introduction

After a severe traumatic experience, some individuals may develop posttraumatic stress disorder (PTSD) (1). PTSD symptoms include intrusive trauma-recollections, avoidance behaviors, negative alterations in cognition and mood, and hyperarousal symptoms (1). Although various evidence-based treatments-including psychotherapy (2, 4) and pharmacotherapy (6, 8) -are available for PTSD (9), current options are not efficacious for all patients: dropout rates (∼16% for psychological therapies) (10), posttreatment symptoms (11), relapse (23.8% following CBT) (12), and treatment resistance (nonresponse up to 50%) (13, 14) are considerable. This clearly illustrates the need for more effective, neurobiologically informed, treatments for PTSD.

Clinical and translational neuroscience have generated models of PTSD psychopathology that highlight abnormalities in the neurocircuitries underling fear learning, threat detection, emotion regulation and context processing (in fear and reward) (15, 16), yet full understanding of PTSD psychopathology, which is essential for the identification of novel therapeutic targets, is still limited (17, 18). Many neurobiological models place alterations in learning and memory of stressful/fearful information -and their subsequent effects on emotional functioning -at a central position in PTSD pathology (e.g. (19–22)). Indeed, this framework can explain aspects of PTSD pathology (21) and the mechanisms of action in psychotherapy (23). Yet, it does not incorporate the impairments in learning and memory of neutral information, which are consistently observed in neuropsychological meta-analyses on PTSD (e.g. (24–26)). These impairments are nevertheless an important part of PTSD’s clinical reality, as they negatively affect treatment responses to psychotherapy (27) and patients’ life satisfaction (28), as well as social and occupational functioning (29).

Together the evidence above illustrates that 1) learning and memory processes play a central role in PTSD pathology, and 2) abnormalities in the underlying neurobiological systems are likely to affect the processing of both emotional/fearful and neutral information, which in turn can influence treatment efficacy. To date, though, there is no comprehensive systematic literature overview available that evaluates PTSD patient’s abilities to learn and memorize neutral and emotionally valenced information together. To fill this gap, we performed a systematic review and meta-analysis to provide an up-to-date overview of current knowledge on learning and memory in PTSD, assessed with behavioral tasks including neutral, emotional, and fearful information (plus fear extinction). Preclinical studies were also evaluated, in addition to clinical studies, as animal models can offer valuable insights in PTSD’s neurobiology and foster drug-development when their phenotype aligns -at least partly-with clinical reality (30–32). Therefore, our primary aim was to evaluate the cross-valence mnemonic performance of i) PTSD patients and ii) animals in PTSD models, compared to their appropriate healthy control group. Following the PTSD psychopathology models (20–22), neuropsychological evidence (24–26), and systematic observations in animal models of PTSD (32, 33), we hypothesize that, cross-species, emotional/fearful learning and memory are enhanced, while fear extinction and neutral learning and memory are impaired in PTSD.

Importantly, PTSD’s heterogeneous nature leads to a diverse patient group (34, 35), and learning and memory processes are especially prone to inter-individual differences (36–38). To address this, our secondary aim was to explore which variables explain variation (heterogeneity) within the clinical and preclinical data. The identification of factors that explain individual variation in learning and memory processes in PTSD is important, as it has been hypothesized that inter-individual differences e.g. in response to traumatic stress play an important role in PTSD psychopathology (39) and resilience (40, 41). It is highly likely that the identification of underlying abnormalities in specific PTSD phenotypes will promote personalized precision medicine for PTSD in the future (42).

## 2. Methods and Materials

This preregistered project (PROSPERO CRD42017062309) (43) is performed in accordance with the PRISMA (44), MOOSE (45), SYRCLE (46, 47) RRIVE (48) guidelines.

### 2.1 Search strategy & Screening

Materials, data, and R-code used for literature search, screening, data extraction and meta-analysis are available via Open Science Framework (OSF; https://osf.io/8ypm5/). A comprehensive literature search on PTSD + Learning and Memory^1^ + Behavior was conducted in the electronic PubMed database (final search on May 22^th^ 2020). The two specific search strings for clinical and preclinical data are provided in Appendix A1. Retrieved articles were independently screened by MS and EG for eligibility against a priori defined inclusion criteria (Appendix A2): 1) PTSD group / model, 2) healthy control group, 3) experimental study, 4) adults, 5) learning/memory/fear-conditioning task, 6) behavioral memory measure, 7) post-trauma memory measure, 8) article in English and essential data available. Discrepancies were discussed until consensus was reached. If eligibility could not be determined based on title and abstract, full-text articles were checked.

### 2.2 Data extraction & study quality assessment

A priori defined data from eligible studies was extracted by one researcher, and independently checked by another. The data extraction codebook is provided as Appendix A3 and included details about 1) publication (author, year), 2) sample (e.g. n, age, sex), 3) trauma and PTSD (e.g. trauma type, time since trauma), 4) learning/memory task (e.g. task, measure), 5) memory performance (mean, SD/SEM).

Data that was exclusively presented in graphs was digitalized with Plot Digitizer (49) and authors were not contacted for missing or additional data. Missing values were included in the data and processed as described in section 2.3.2.

Study quality and risk of bias were assessed with an adapted version of the Newcastle-Ottawa case-control Scale (NOS) (3) (see Appendix A4) in the clinical case-control studies, and with SYRCLE’s risk of bias tool (5) in the experimental preclinical studies. On both scales, unreported details were scored as an unclear risk of bias.

### 2.3 Meta-analysis

The analytic strategy is based on earlier work of our group (50, 51) and performed with α = .05 in R version 4.0.3 (52), with the use of packages dplyr (53), purr (54), tidyr (55), osfr (56), metafor (57), metaforest (58), caret (59), metacart (60), ggplot2 (61), ggpubr (62), gridExtra (63), Gmisc (64), viridis (65), and arsenal (66). As effect size we calculated the standardized mean difference Hedge’s G (57). Clinical and preclinical data were always analyzed as separate datasets.

#### 2.3.1 Random-effects meta-regression: valence x phase

To answer the primary research question, the overall effect size per valence type (neutral, emotional, fear and trauma) and phase (learning, memory and extinction) was estimated with a nested random-effects model with restricted maximum likelihood estimation and valence x phase as moderator, as variation between studies (heterogeneity) was expected (67). The estimation was nested within studies and independent PTSD groups (experimental groups). Combinations of valence and phase that were not present in the data were excluded from the model (e.g. neutral + extinction); levels of categorical variables with <4 studies were also excluded (68). P-values were Bonferroni corrected within the clinical and preclinical dataset.

Cochrane Q-test (57) and the I^2^-statistic were used to asses heterogeneity. I^2^ of 25%, 50% and 75% represent respectively low, moderate and high levels of heterogeneity (69). Rosenthal’s fail-safe N (70) was calculated for each valence x phase level in the models, to evaluate the robustness of the estimated effects. Egger’s regression (71) was used to asses funnel plot asymmetry as an index for publication bias. The potential influence of 1) study quality, 2) outliers and influential cases (7), and 3) comparison type was evaluated with a sensitivity analysis. To evaluate the influence of study quality, the scores on NOS (for clinical data) and SYRCLE’s risk of bias tool (for preclinical data) were combined into summary risk of bias scores (yes = 0; unclear = 0.5; no = 1), where higher scores represent more risk of bias.

#### 2.3.2 Exploratory analysis

The sources of variation (heterogeneity) within the clinical and preclinical subgroup were explored with a two-step data-driven analysis. Missing values (<1/3 missing) in ‘sex’ and ‘time since traum’ were replaced by the most prevalent category and median value, respectively. No missing values were present in the other variables.

First, potential moderators of the effect sizes were ranked based on their permuted variable importance in MetaForest, a random forest-based meta-analysis (72). The 10-fold cross-validated random-forests (500 trees) were tuned for minimal RMSE in clinical (fixed weighting, 2 candidate moderators at each split, minimum node size of 6) and preclinical (random weighting, 6 candidate moderators at each split, minimum node size of 2) data separately. Models showed good convergence (Figure S4-S5). The predicted effect size by different levels of a specific moderator -when all other moderators are kept constant-were explored via partial dependence (PD) plots (72, 73).

Next, potential interactions between moderators were explored by fitting a tree-based random-effects meta-CART algorithm with look-ahead strategy to the datasets (pruning parameter c=0.5, maximum of 10 splits, 10-fold cross-validation) (74). Although tree-based models (like meta-CART) are less stable and more prone to overfitting than random-forest-based models (like MetaForest), meta-CART has an advantage over the ‘black box’ MetaForest in its ability to provide interpretable interactions (72, 74). As advised, to overcome the potential instability of meta-CART, the suggested interactions were explored via PD plots in the MetaForest model (72).

## 3. Results

### 3.1 Study selection and characteristics

After screening of 1661 records, 92 clinical (6732 humans) and 182 preclinical (6834 animals) studies were included in the meta-analysis (Figure 1). Characteristics of these studies are provided in Table S4-S6. The independent clinical PTSD groups represented civilians (52%) and veterans (48%) and were mostly of mixed gender (58%) and middle-aged (56%). Most independent preclinical PTSD groups contained rats (83%), males (94%) and young adults (88%). Most clinical PTSD groups were compared to trauma-exposed (61%) or non-exposed (37%) controls, while the majority of preclinical PTSD groups were defined as ‘trauma-exposed’ and compared to non-exposed controls (93%). Cued tasks (94%) and neutral valenced information (64%) were mostly used in clinical groups, while contextual tasks (70%) and fear (39%) and trauma-related (46%) information were mostly assessed in preclinical groups.

**Figure 1.**
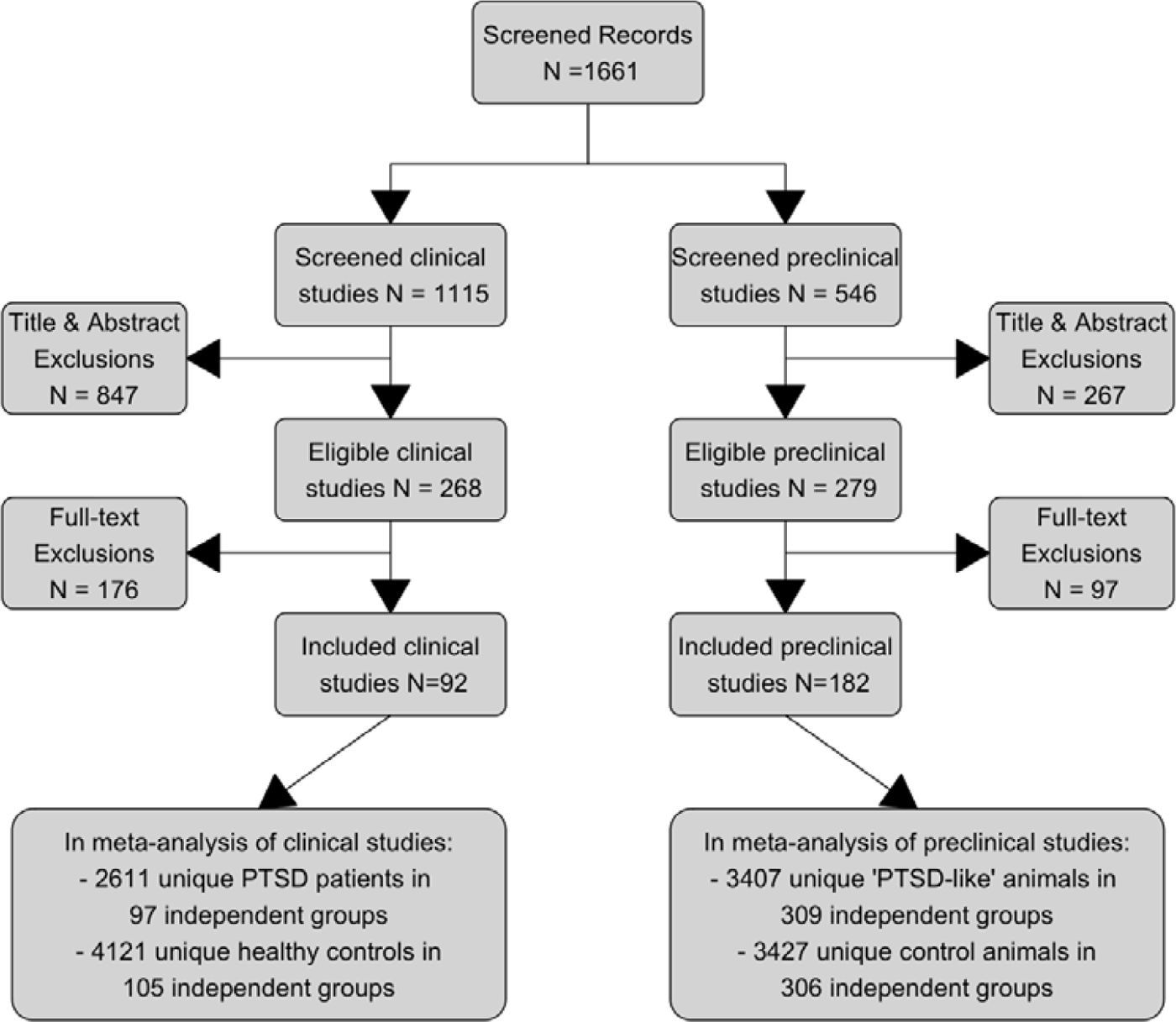
Flowchart.

More than 70% of the clinical studies reported low risk of bias on all NOS items, except bias due to non-response rates during recruitment (low risk only reported in 10% of the studies). In preclinical studies, reporting was less adequate: no studies reported on all SYRCLE’s items (Figure S1). Risk of bias due to (non-)random housing (100%), (non-)random outcome assessment (100%), and allocation concealment (97%) was unclear in almost all preclinical studies. Most preclinical studies were at high risk of bias due to unblinded experimenters (65%), but at low risk of bias due to (equal) baseline characteristics (98%) and blinded outcome assessment (59%).

### 3.2 Effect of PTSD on neutral and emotional learning, memory, and extinction

The random-effects meta-regression on clinical data (Figure 2.A; Table S7) showed that PTSD patients have an impaired ability to learn neutral information (Hedge’s G = −0.667, p <.001), remember neutral (Hedge’s G = −0.544, p <.001), and emotional material (Hedge’s G = −0.655, p <.001), and extinguish fearful information (Hedge’s G = −0.804, p<.001), compared to healthy controls. Fear learning did not differ significantly between PTSD patients and healthy controls (Hedge’s G = −0.200, p=1). The effect of PTSD on trauma learning (1 study), fear memory (1 study) and trauma memory (2 studies) could not be estimated reliably in the clinical dataset, due to insufficient number of studies (<4). In animal models of PTSD the impairments in neutral learning (Hedge’s G = −1.304, p <.001) and neutral memory (Hedge’s G = −1.291, p <.001) were also present (Figure 2.B; Table S8). Moreover, enhanced fear learning (Hedge’s G = 0.435, p= .034), stronger memory for fear (Hedge’s G = 0.812, p <.001) and especially trauma-related (Hedge’s G = 1.877, p <.001) material, was observed compared to controls (fear vs trauma memory: difference in Hedge’s G = 1.065, p<.001). To explore how this relates to the reduction in patient’s emotional memory performance, we estimated the effect sizes of the three clinical studies (PTSD patients: n=52; healthy controls: n=67) that measured fear and trauma memory. This explorative analysis revealed positive effect sizes which might indicate that trauma (Hedge’s G=0.357; p=.162) and fear (Hedge’s G=0.425; p=.252) memory is also enhanced in PTSD patients, but these estimations should be interpreted with caution as they are based on less than the recommended 4 studies per subgroup (68). Finally, as in humans the preclinical data shows reduced fear extinction (Hedge’s G = −0.741, p <.001); interestingly, extinction of trauma-related material (Hedge’s G = −2.190, p<.001) was more impaired than other forms of fear extinction (difference in Hedge’s G = −1.449, p<.001).

**Figure 2.**
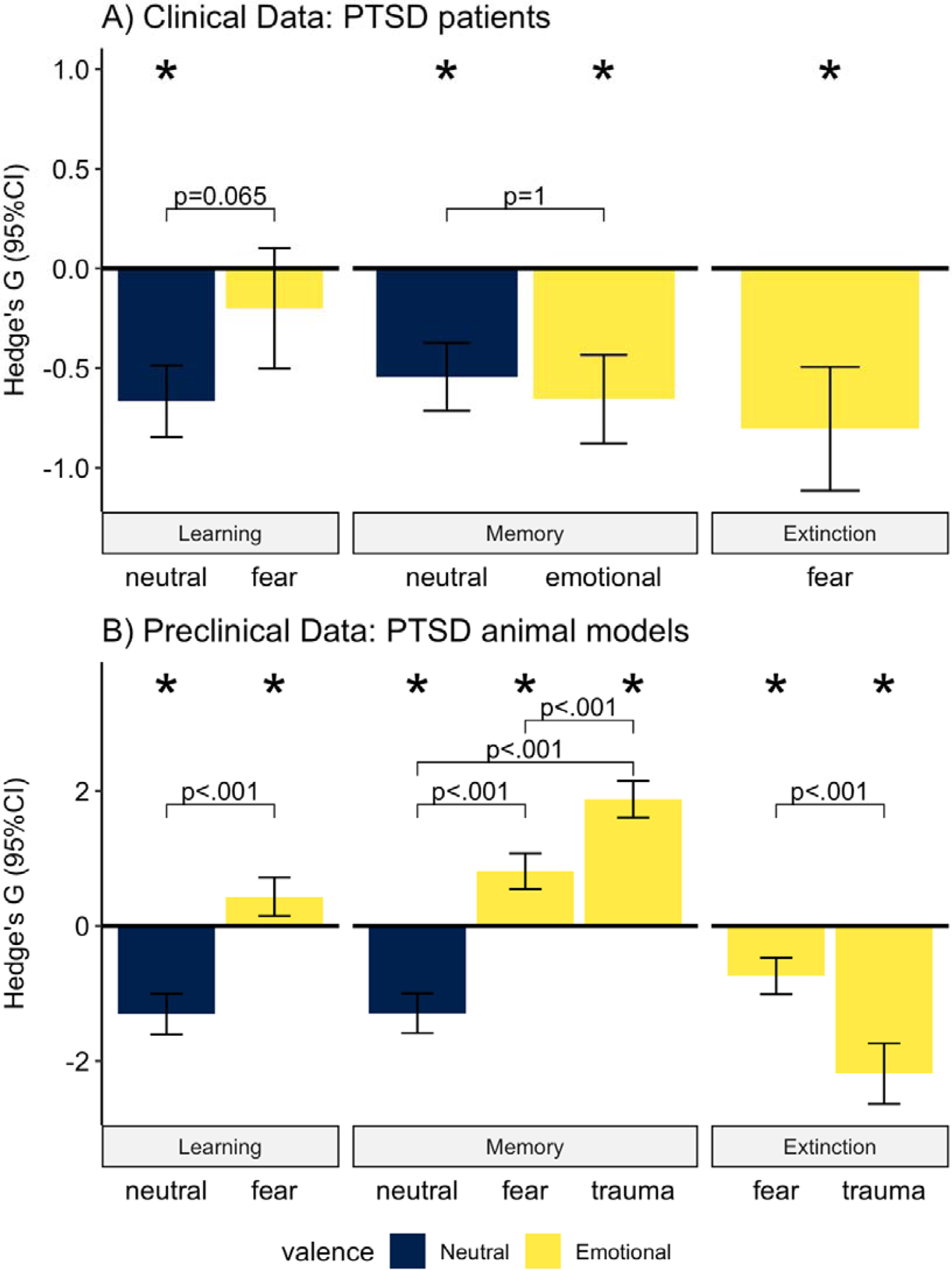
Meta-regression: cross-valence mnemonic performance in clinical and preclinical data. The standardized mean difference Hedge’s G and 95% confidence intervals. Positive effect sizes indicate improved performance in PTSD, negative effect size indicates reduced performance in PTSD. * Indicates effect size was significantly different from 0 (Bonferroni corrected P < 0.05).

### 3.3 Robustness of the effect

Substantial heterogeneity was observed in both clinical (Q(542) = 7242.000, p < .001; I^2^ = 83.97, 75,42% between study variance, 8.55 % within study variance) and preclinical data (Q(1082) = 5762.943, p < .001; I^2^ = 88.60, 75.56% between study variance, 13.03% within study variance). Qualitative evaluation of funnel plot asymmetry suggests some publication bias, which was confirmed by Egger’s regression in clinical (Figure S2), not preclinical (Figure S3), data. Yet Rosenthal’s fail-safe N analyses suggest that this is unlikely to influence interpretation of the clinical (Table S9) or preclinical (Table S10) results. Sensitivity analysis confirmed that study quality was not a significant moderator of the overall effect in clinical (Q(1) = 0.062, p= 0.804) and preclinical (Q(1) = 0.089, p = 0.766) data. Nor did exclusion of influential cases and outliers change the clinical (Table S11) or preclinical (Table S18) results. The influence of comparison type remains partly inconclusive, due to insufficient data for some combinations of phase and valence. For most categories with sufficient data (≥ 4 studies) findings of the main analysis were also observed in each comparison type (Table 1), except for enhanced fear memory in preclinical data: which was not present when trauma-exposed controls were compared to animals with PTSD like behavior.

**Table 1.** Summary sensitivity analysis by comparison type.

| Comparison type | Clinical studies | Predclinical studies |
| --- | --- | --- |
| **Non-trauma-exposed controls vs PTSD group** | **Similar** results as main analysis for categories with $\geq 4$ studies (Table S12-S13) | **Inconclusive:** Limited data available in some categories. Only $\geq 4$ studies on trauma memory: in this category similar results as in main analysis (Table S21-S22) |
| **Non-trauma-exposed controls vs trauma-exposed controls** | **Inconclusive:** Limited data available, no categories with $\geq 4$ studies (Table S14-S15) | **Similar** results as main analysis for categories with $\geq 4$ studies (Table S19-S20) |
| **Trauma-exposed controls vs PTSD group** | **Similar** results as main analysis for categories with $\geq 4$ studies (Table S16-S17) | **Inconclusive:** Limited data available in some categories. Only $\geq 4$ studies on fear and trauma memory: similar results as in main analysis were observed for trauma memory. In contrast to the main analysis no significant difference between groups was observed for fear memory (4 studies). (Table S23-S24) |
| * The Appendix B5 provides an overview of the available data and sensitivity analysis results per comparison type for clinical studies (Table S12-S17) and preclinical studies (Table S19-S24). |  |  |

### 3.4 Potential moderators in clinical studies

Together the variables in the MetaForest model explained only 8% of the variance in effect sizes in the clinical dataset (Rcv^2^ [SD]= 0.081 [0.106]). The ranking of the moderators is shown in Figure 3. Information type, sample and phase were selected as the most important variables, but the relatively low variable importance scores (Figure 3) and 8% total variance explained (Rcv^2^) do not suggest strong moderation. Indeed, the follow-up PD plots suggest that performance is generally impaired (Hedge’s G ∼ 0.5) across all levels of the evaluated moderators (Figure S6). In the absence of strong moderators, no further meta-CART analysis was performed on the clinical dataset. Together with the impaired performance of PTSD patients in all categories of the meta-regression, these findings suggest a general impairment in learning, memory, and extinction in PTSD patients, as far as evaluated. Of note, the influence of cue/context remains inconclusive due to the limited variation in the dataset (94 % cued tasks).

**Figure 3.**
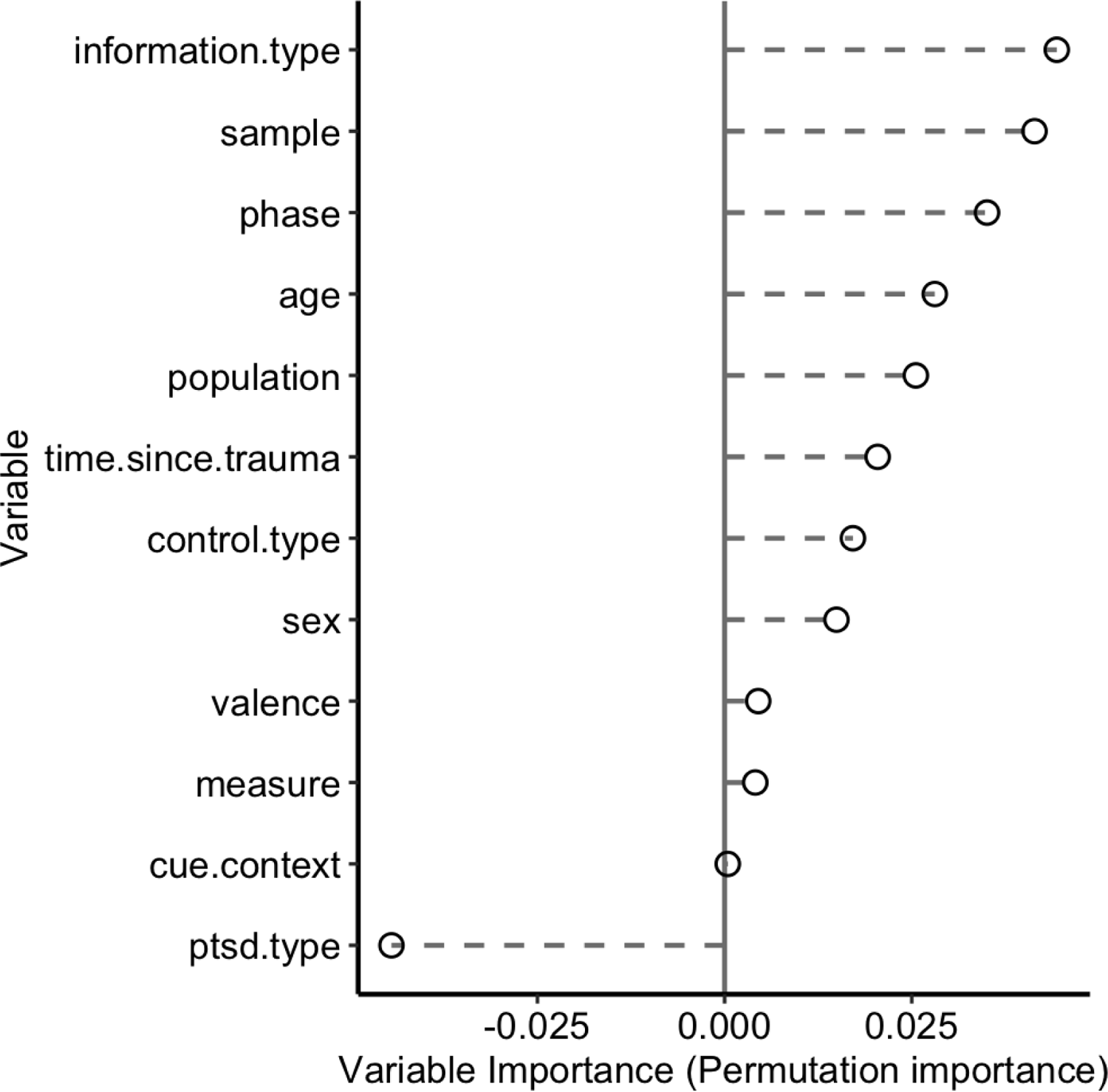
Clinical MetaForest variable importance. Relative importance of potential moderators based on ‘permuted variable importance’ in the random forest-based meta-analyses on clinical data. In total, 8% variance in effect sizes was explained by the MetaForest model. Indeed, the low variable importance score do not suggest strong moderation by any of the estimated variables.

### 3.5 Potential moderators in preclinical studies

For the preclinical data, 53.4% of the variance in effect sizes was explained by the variables evaluated with MetaForest (Rcv^2^ [Sd]=0.534 [0.096]). This is a considerable amount, and the variable ranking shown in Figure 4 indicates that phase and valence are the most important moderators, followed by information type (i.e. olfactory vs safety vs spatial vs threat vs visual information). Note, these moderators were also included in the meta-regression, which illustrates that the most important factors were evaluated in this analysis. Indeed, the follow-up PD plots (Figure S7) of these variables correspond with the results of the meta-regression.

**Figure 4.**
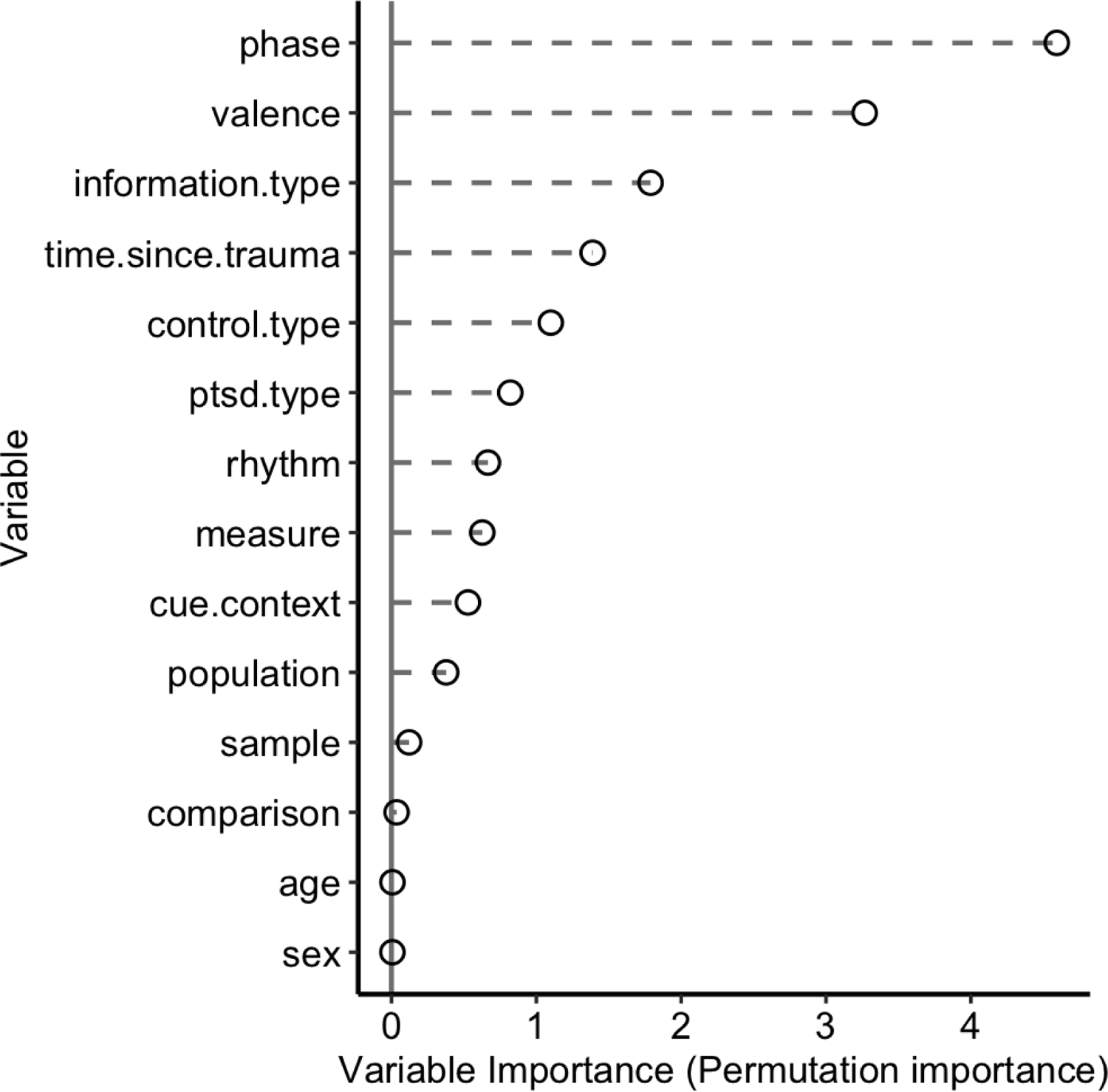
Preclinical MetaForest variable importance. Relative importance of potential moderators based on ‘permuted variable importance’ in the random forest-based meta-analyses on pre-clinical data. In total, the MetaForest model explained 53.4 % of variance in effect sizes. Phase and valence are selected as the most important variables, there effects were also evaluated in the meta-regression.

Meta-CART suggested that the effect sizes in preclinical data were influenced by phase and PTSD type, each in interaction with information type (figure S8). Exploration of these interactions in MetaForest PD plots (figure S9) only provides evidence for a *phase* x *information* type interaction, largely in line with the meta-regression analysis.

## 4. Discussion

Here we report the first comprehensive meta-analysis on learning, memory, and extinction of neutral and emotional (including fearful and trauma-related) information in PTSD patients and animal models of PTSD. The results confirmed the hypothesis that neutral learning/memory and fear extinction are cross-species impaired in PTSD, but the expected stronger fear memory in PTSD was only observed in preclinical studies (PTSD patients showed impaired emotional memory).

Clinical and preclinical studies differed in some characteristics. Clinical studies mostly investigated older patients of mixed gender, while preclinical studies typically included younger, male animals. Stronger effect sizes were observed in preclinical studies (likely due to their controlled nature) and overall reporting on potential risks of bias was better in clinical studies. Importantly, animals exposed to trauma were typically considered to represent ‘the PTSD group’ in most preclinical studies. This is an inaccurate conceptualization of PTSD, as clinical studies showed that only a minority of trauma-exposed individuals actually develop PTSD, which was confirmed in those animals studies that addressed the issue (reviewed in (30)). Unfortunately, insufficient data was available to quantify the influence of this experimental difference, but future preclinical studies should pay attention to this inconsistency.

Partly as expected, learning, memory, and extinction were impaired in PTSD patients. This impairment is strong for both neutral as well as emotionally valenced material. No strong moderators of these effects were identified, suggesting a general impairment of learning and memory processes in PTSD patients. This contrasts, for example, with earlier reports on the influence of sex on fear conditioning (75), or on HPA-axis function in PTSD (76). Like PTSD patients, animal in PTSD models showed impaired neutral learning/memory and fear extinction (especially for trauma-related information). The reduced extinction in preclinical data might be hampered by strong fear (and mostly trauma) memories that compete with fear expression during extinction learning (77–79). PTSD patients could show a similar phenotype, but definite conclusions await more studies. Phase and valence were the strongest moderators of performance in animal models (pointing towards limited influence of age, sex, PTSD-model, species, strain, etc.). Note, the apparent lack of importance of age and sex in the preclinical dataset can also be due to limited variation in these variables (i.e. mostly young adult and male animals). Together the preclinical and (as far as available) clinical data seem to indicate that PTSD affects neutral and emotional on the one hand versus fear/trauma memory on the other hand in opposite directions.

### 4.1 Impaired fear extinction in PTSD

Impaired fear extinction is a strong phenotype in PTSD patients and animal models, as evidenced by large effect sizes, despite substantial heterogeneity in the data. Animal models highlight that extinction of trauma-related information is particularly impaired. This confirms current neurobiological models of PTSD (15, 21) and justifies that extinction is the prime target of exposure-based psychotherapies for PTSD (23, 80). However, the observation that impaired extinction is not limited to trauma-related information might also indicate that the neurobiological mechanisms underlying the extinction process itself does not function optimally in PTSD patients. There is even some evidence that this is a pre-existing trait which makes these subjects vulnerable to the development of PTSD in the face of trauma (81, 82). Indeed, abnormalities in brain areas involved-amygdala, hippocampus, and prefrontal cortex-have been observed in PTSD (19). Perhaps, this explains why exposure-treatments that rely on the patients ‘existing’ extinction abilities can be less effective for some patients. Indeed, extinction abilities vary between individuals (83) and some patients might benefit from complementary therapies that boost extinction (84). Various psychological, behavioral, brain-stimulation, and psychopharmacological interventions hold the potential to augment extinction (84). Interestingly, there is a range of possible neurobiological targets, including synaptic plasticity (85, 86), prefrontal cortex-amygdala / hippocampus connectivity (87–90), and several neurotransmitter systems (91, 92), including serotonin, dopamine (93–96), noradrenalin (97–99), choline (100), glutamate, GABA, (endo)cannabinoid (101–103), glucocorticoid (104–106), and others. Although successful translation of single-target interventions into clinical practice is still limited (84, 106), this range opens possibilities for the development of multi-target approaches, tailored to the patient-specific neurobiological abnormalities that underlie impaired extinction (84, 91). Our results indicate that preclinical studies can accurately model this clinical phenotype and serve to develop new therapies.

### 4.2 Impaired neutral learning and memory in PTSD

Reduced ability to learn and memorize neutral information were another strong phenotype in both PTSD patients and animal models. This phenotype should not be overlooked in PTSD research and clinical practice, as it is just as prevalent as impaired extinction and can pose a substantial burden on PTSD patients’ daily functioning and treatment response (107, 108). Moreover, a prospective study showed that deficits in neutral learning and memory contribute to PTSD vulnerability (109). One can speculate that this phenotype hampers the discrimination between safe and neutral events (110), thereby contributing to impaired safety learning in PTSD patients (111–113). Interestingly, it has been found that psychotherapy can improve verbal memory in PTSD (114), which might be explained by its enhancing effects on hippocampal functioning (115) and changes in FKBP5 expression (116). To improve clinical practice, novel (complementary) treatments should target this phenotype directly, for example via 1) behavioral interventions - such as targeted memory reactivation (117, 118), behavioral tagging (118, 119), reconsolidation updating (118) and reminders (120) - that tap into endogenous encoding and retrieval processes; 2) cognitive training that enhances learning and memory strategies (121, 122); 3) physical exercise - like cardiovascular exercise (123–126) and balance training (127) - that stimulates the hippocampal memory system; 4) sleep interventions that enhance slow-wave sleep (128); 5) neurofeedback training that improves prefrontal cortex-hippocampus connectivity (129); or 6) pharmacotherapy that targets neurotransmitter systems (e.g. serotonin (130), dopamine (131), choline (132, 133)) or modulates neuronal processes (e.g. neurogenesis (134), neuro-inflammation (134), or neuronal damage (126)). Importantly for future neurobiological research and drug-development, our results show that this phenotype is also present in animal models of PTSD.

### 4.3 Differences in emotional and fearful/trauma memory

Contrary to fear extinction and neutral memory formation, our meta-analysis suggests that PTSD might have opposing effects on emotional (impaired in clinical data) and fearful/trauma memory (improved in preclinical data) in clinical versus preclinical studies. One explanation may be linked to the stress system. Thus, in humans emotional memory tasks are unlikely to trigger activation of the HPA-axis (e.g. (135)), while preclinical fear conditioning tasks most certainly do (e.g. (136)). Our findings suggest that PTSD (or trauma exposure) related changes in HPA-axis functioning benefit memory for fearful information - as corticosteroids can enhance memory formation (137)-, at the cost of neutral and slightly emotional information. This conclusion should be drawn with caution, though, as studies on HPA-axis alterations in PTSD yield mixed results (76) and there was insufficient data available on fearful/trauma memory in patients and emotional memory in animal models for the current meta-analysis. Another explanation could be that animal models and PTSD patients show an opposing phenotype (i.e. impaired vs enhanced emotional learning in PTSD). If present, this would be problematic for drug-development research, as agents that reduce (enhanced) emotional learning in animal models of PTSD would inevitably fail to improve (impaired) emotional learning in PTSD patients.

### 4.4 Strengths and limitations

The large cross-species dataset (274 studies), and integrated hypothesis-driven meta-regression with state-of-the-art heterogeneity exploration via random-forest and tree-based models are strengths of this meta-analysis. To improve cross-species comparability, only behavioral measures of learning and memory were included, which might limit the generalizability of our findings to, for example, physiological, neuroimaging or self-report measures.

### 4.5 Conclusion

All in all, this meta-analysis shows that both impaired neutral learning/memory and fear extinction are two strong clinical phenotypes of PTSD, that can be accurately modeled in preclinical studies. Novel PTSD treatments could target these phenotypes and benefit from animal models to unravel the underlying neurobiology and foster drug-development. In addition, future research should elaborate on the origin of potential differences between emotional and fear/trauma memory in PTSD across species. Until this issue is resolved, we do not recommend to use animal models for drug-development that targets emotional/fearful memory in PTSD.

## Data Availability

Materials, data, and R-code used for literature search, screening, data extraction and meta-analysis are available via Open Science Framework (OSF; https://osf.io/8ypm5/).

## Acknowledgments

We thank Lara Oblak and Sophie Heijnen for their help with screening and data-extraction. We also thank Valeria Bonapersona for her statistical advice and valuable discussions. This work is supported by the Dutch Ministry of Defense. MS is supported by a personal grant which is part of the Graduate Program (project #022.003.003) of The Netherlands Organization of Scientific Research NWO. MS and MJ were supported by the Consortium on Individual Development (CID), which is funded through the Gravitation program of the Dutch Ministry of Education, Culture, and Science and Netherlands Organization for Scientific Research (project #024.001.003). The funders had no role in study design, data collection and analysis, decision to publish, or preparation of the manuscript. This manuscript is shared as preprint on medRxiv.

## Footnotes

1 As the initial search performed on PTSD + cognition + behavior yielded mostly results on learning and memory, the research focus and subsequent searches were limited to this cognitive domain.

