## Appendix for "Impaired learning, memory, and extinction in posttraumatic stress disorder: translational meta-analysis of clinical and preclinical studies"

### SUPPLEMENTAL INFORMATION (APPENDIX)

### A. Supplemental methods

#### A1. PubMed search strings

General structure of the search filters is provided in the table S1 below. The complete clinal and preclinical search strings are provided on OSF via https://osf.io/v3zn2/ and https://osf.io/928ue/ respectively.

| Table S1. **General structure of Pubmed search filters** |
|---|
| **Learning & memory: Clinical + Preclinical** |
| (“learn”[tiab] OR “learning”[tiab] OR “memories”[tiab] OR “memory”[tiab] OR “Memory, Episodic“[Mesh] OR “Memory, Short-Term“[Mesh] OR “Immediate Recall“[tiab] OR “Immediate Recalls“[tiab] OR “Learning“[MeSH] OR “Association“[Mesh] OR “Association learning“[MeSH] OR “Avoidance Learning“[MeSH] OR “Conditioning (Psychology)“[Mesh] OR “Conditioning“[tiab] OR “Conditioning, Classical“[Mesh] OR “Conditioned Reflex“[tiab] OR “Conditioning, Eyelid“[Mesh] OR “Conditioning, Operant“[Mesh] OR “Cues“[MeSH] OR “Discrimination Learning“[MeSH] OR “Generalization (Psychology)“[MeSH] OR “Fear Generalization“[tiab] OR “Generalization, Response“[Mesh] OR “Generalization, Stimulus“[Mesh] OR “Stimulus Generalization“[tiab] OR “Memory“[Mesh] OR “Memory, Long-Term“[Mesh] OR “Memory Consolidation“[Mesh] OR “Consolidation“[tiab] OR “Mental Recall“[Mesh] OR “Mental Recall“[tiab] OR “Familiarity“[tiab] OR “Retention (Psychology)“[Mesh] OR “Retention“[tiab] OR “Spatial Memory“[Mesh] OR “Reinforcement (Psychology)“[Mesh] OR “Reinforcement“[tiab] OR “Extinction, Psychological“[Mesh] OR “Extinction“[tiab] OR “Reversal Learning“[Mesh] OR “Set (Psychology)“[Mesh] OR “Spatial Learning“[Mesh] OR “Maze Learning“[Mesh] OR “Verbal Learning“[Mesh] OR “Paired-Associate Learning“[Mesh] OR “Serial Learning“[Mesh] OR “trauma cue freezing”[tiab]) |
| **PTSD: Clinical + Preclinical** |
| ("Trauma and Stressor Related Disorders"[Mesh] OR "Trauma and Stressor Related Disorders"[tiab] OR "Stress Disorders, Traumatic"[Mesh] OR "Psychological Trauma"[Mesh] OR "Stress Disorders, Post-Traumatic"[Mesh] OR “Traumatic Stress Disorder”[tiab] OR “Traumatic Stress Disorders”[tiab] OR “Combat Stress Disorders”[tiab] OR “PTSD”[tiab] OR “Posttraumatic Stress Disorders”[tiab] OR “Posttraumatic Stress Disorder”[tiab] OR "Combat Disorders"[Mesh] OR "Combat Disorders"[tiab] OR "Combat Disorder"[tiab] OR “Combat Neurosis"[tiab] OR “Combat Neuroses"[tiab] OR “Shell Shock"[tiab] OR “War Neurosis"[tiab] OR “War Neuroses"[tiab] OR “Combat Stress Disorder"[tiab] OR “Post-Traumatic Neuroses”[tiab] OR “Posttraumatic Neuroses”[tiab] OR “traumatic stress symptoms”[tiab]) |
| **PTSD: addition Preclinical** |
| “model”[tiab] |
| **Behavior: Clinical + Preclinical** |
| (“discrimination”[tiab] OR “freezing”[tiab] OR “false alarm rate”[tiab] OR “hit rate”[tiab] OR “startle”[tiab] OR “encoding”[tiab] OR “retrieval”[tiab] OR “recall”[tiab] OR “recognition”[tiab] OR “learning task”[tiab] OR “memory task”[tiab] OR “learning test”[tiab] OR “memory test”[tiab] OR “fear conditioning”[tiab] OR “conditioned avoidance”[tiab] OR “memory scale”[tiab] OR “memory measure*”[tiab] OR “maze”[tiab] OR “foot shock”[tiab] OR ( (“neuropsychological“[tiab] OR “neurocognitive”[tiab] OR “neuropsychology”[tiab] OR “cognitive”[tiab] OR “behavior*"[tiab] OR “Behaviour*”[tiab]) AND (“Task*”[tiab] OR “Test*”[tiab] OR “assessment”[tiab] OR “measure*”[tiab] OR “score*”[tiab] OR “performance*”[tiab] OR “response*”[tiab]) OR “profile*”[tiab]) ) |

| Subjects: Preclinical | Subjects: Clinical |
|---|---|
| <SYRCLE's animal filter> | ( ("Patients"[Mesh] OR "Patient*"[tiab] OR "Client*"[tiab] OR "Veterans"[Mesh] OR "veteran*"[tiab] OR "survivor*"[tiab]) NOT ("mice"[tiab] OR "rats"[tiab] OR "rat"[tiab] OR "mouse"[tiab] OR "animal model"[tiab]) ) |
| **Only experimental: Clinical + Preclinical** | |
| NOT ("case"[ti] OR "meta-analys*"[ti] OR "review"[ti] OR Review [Publication Type] OR systematic[sb] OR "meta-analysis"[Publication Type] OR "Case Reports"[Publication Type]) | |

#### A2. Inclusion / Exclusion criteria

Table S2. Inclusion and exclusion criteria

| | Inclusion criteria | | | Exclusion criteria | | |
|---|---|---|---|---|---|---|
| | Description | Operationalization (if applicable and not exhaustive) | | Description | Operationalization (if applicable and not exhaustive) | |
| | | Clinical | Preclinical | | Clinical | Preclinical |
| 1 | **PTSD group / model** | valid diagnosis by clinician (e.g. psychologist, psychiatrist, etc.) or via structured interview (e.g. CAPS, SCID, DSM, DIAX/M-CIDI, MINI, Mini-DIPS, F-DIPS, ADIS-R, etc.) | valid PTSD models according to (1,2); footshock >= 0.8mA (2); re-stress or SR models (not considered chronic PTSD) | **no PTSD group or model: chronic PTSD [1] / Early life stress / (mild) traumatic brain injury / burn-injury** | correlations with symptoms; no PTSD-only group (comorbidity); partical/subclinical PTSD; chronic PTSD [1]; complex PTSD; lifetime PTSD (not current); PTSD based on self-report questionnaire cut-off (e.g. PCL-C, PCL-M, etc.) | chronic ptsd: trauma duration > 1 day; pain models (e.g. pain threshold shock); genetically modified animals (e.g. with increased stress-suseptibility) without additional exposure to a traumatic sitmulus (i.e. absence of PTSD Criterium A) |
| 2 | **healthy control group** | | | **no healthy control or no explicit comparison** | (m)TBI; psychiatric or neurological illnesse (e.g clinical cognitive impairement, stroke); chronic physical illness | |

[1] Clinical and preclinical differ with respect to the chronic-criterium. Although chronic PTSD is not the same as chronic trauma, both were exclude as they likely affect behavior differently then acute PTSD/Trauma. Borghans (2015) does not differentiate between acute and chronic preclinical models, Flandreau (2017) does. We followed Flandreau, which is a more critical review compared to Borghans' observational review.

| | | | | | | |
|---|---|---|---|---|---|---|
| | | | | between PTSD-only and healthy controls | | |
| 3 | experimental study | | | no experimental study | case reports; reviews | reviews |
| 4 | adolescent/adult subjects | 15 – 65 years | 6 weeks - 1 year [2]; 'adult' | no adolescents/adults (outside range) or age unclear / non-reported | childeren & elderly [3] | |
| 5 | learning / memory / fear conditioning tasks | | escape measures in shuttlebox or licht-dark box if the trauma (e.g. shock) was in this box (which was considered avoidance due to trauma memory); (contextual) fear conditioning; trauma (cue/context) memory; etc. | no learning / memory / fear conditioning tasks (other domains) | tasks that involve:<br>1. working memory (e.g. (dual)sternberg task; etc.)<br>2. cognitive control (inhibition)<br>3. reward (e.g. punishment learning with monitary loss; beautiful faces; Probabilistic classification task; etc.)<br>4. short tem reading comprehension tasks (= language processing, not memory) | tasks that involve:<br>1. reward (e.g. place to cue set-shifting (cue to location of reward); conditioned place preferance; press levers for food (operant learning); etc.)<br>2. depression/learned helplessness (e.g. escape measures in shutttle box if trauma (usually footshock) was NOT in this box; forced swim test);<br>3. general anxiety without learned component (e.g. 5 |

[2] if only weight range was provided, the average is used to deterime age of animals (http://www.arc.wa.gov.au/?page_id=125; https://www.criver.com/products-services/find-model/wistar-igs-rat?region=3611; https://www.criver.com/products-services/find-model/sas-sprague-dawley-rat?region=3651; https://www.criver.com/products-services/find-model/lewis-rat?region=3651). Ages were rounded to 0-decimal numbers (i.e. >= 5.5 weeks were included).

[3] For one clinical study (PMID 17008144), with two PTSD groups with average ages of 67 and 72 years, the 67-aged group was included and the 72-aged group was excluded (following the liberal approach to age-weight transformations for animal studies).

| | | | | |
|---|---|---|---|---|
| | | | 5. performance validity tests (Word Memory Task, Green Word Memory Test); symptom validity tests<br>6. emotional cognitive (schema) bias (e.g. episodic future thinking task (= "imagin an event in the future that will give you a certian feeling"); future-based Autobiographical Memory Test; (trauma) word-stem completion tasks; etc.)<br>7. emotion processing/detection (e.g. processing of social signals (threat, anger, anxiety); emotional face-matching task; FPS during trauma stimuli; etc.)<br>8. learning after stress (e.g.TSST)<br>9. general anxiety (e.g. fear generalization) | min after SR freezing in EPM; fear generalization in OF or EPM; sudden silence test; black-light box (see shuttle box above); escape/avoidance or leverpress avoidance (= more motivation to escape, then memory of environment/context); fear sensitization (= innate startle ipv memory measure);<br>4. Modified holeboard (measures working memory, reference strategy, general anxiety, social, but no learned component);<br>5. social interaction test;<br>6. cognitive flexibility (e.g. operant strategy shifting paradigm) |
| 6 | **behavioral measure** | **no behavior** | questionnaire data (e.g. memory complains or problems in daily life); only neuroimaging (e.g. resting-state MRI); autobiografic script-driven imagery ("participants listened to recordings of personal trauma episodes and focused on the evoked | |

| | | | | | | |
|---|---|---|---|---|---|---|
| | | | | | mental images and feelings"); qualitative measures (e.g. open-ended questions on ICU memories); self-report memory (e.g. expectancy or contingency scores FC) | |
| 7 | **post trauma measure (for PTSD cases)** | >= 1 month trauma-measure interval | >= 3 day (~comparible to human 1 month) trauma-measure interval | **pre-trauma tasks (=risk-factor) / shorter interval trauma-measure** | | [4] |
| 8 | **three or more comparisons between PTSD - HC need be present of each category** | | | **categories with less then 3 comparisones were excluded from the dataset** | e.g. prospetive & retrosepctive memory (only 1 study) | |
| 9 | **availability (e.g. essential data online; article in English)** | | | **data not available online (authors were not contacted); article in other language** | | |

[4] Note, if learning was <3 days from trauma, but memory was measured >= 3 day after trauma, memory was included (learning not).

#### A3. Data extraction codebook

https://osf.io/tvne6/

#### A4. Adapted Newcastle-Ottawa case-control Scale (NOS)

##### NEWCASTLE - OTTAWA QUALITY ASSESSMENT SCALE CASE CONTROL STUDIES – adapted for TRACE

Note: A study can be awarded a maximum of one star for each numbered item within the Selection and Exposure categories. A maximum of two stars can be given for Comparability.

**Selection**

1) Is the case definition adequate? / TRACE: validated measure of PTSD?

a) yes, with independent validation ✵ / TRACE: clinical interviews (CAPS/SCID) or diagnosis (Clinician)

b) yes, eg record linkage or based on self-reports / TRACE: self-report questionnaire

c) no description

2) Representativeness of the cases / TRACE: representative PTSD subjects?

a) consecutive or obviously representative series of cases ✵ / TRACE: all eligible PTSD patients in a certain area / subgroup / hospital / clinical / etc.

b) potential for selection biases or not stated

3) Selection of Controls / TRACE: representative control subjects?

a) community controls ✵/ TRACE: controls from the same area / subgroup / etc. as cases

b) hospital controls

c) no description

4) Definition of Controls

a) no history of disease (endpoint) ✵ / TRACE: no (history of) PTSD

b) no description of source

Comparability

1) <u>Comparability of cases and controls on the basis of the design or analysis</u> / TRACE: adjustments for confounders?

a) study controls for ________________ (Select the most important factor.) ✵ / TRACE: yes

b) study controls for any additional factor ✵ (This criteria could be modified to indicate specific control for a second important factor.) / TRACE: -

Exposure / TRACE: learning/memory measures

1) <u>Ascertainment of exposure</u> / TRACE: measurement of learning/memory?

a) secure record (eg surgical records) ✵ / TRACE: behavioural task

b) structured interview where blind to case/control status ✵

c) interview not blinded to case/control status

d) written self report or medical record only

e) no description

2) <u>Same method of ascertainment for cases and controls</u> / TRACE: same task/scoring?

a) yes ✵

b) no

3) <u>Non-Response rate</u>

a) same rate for both groups ✵

b) non respondents described

c) rate different and no designation

### B. Supplemental results

#### B1. Study Characteristics

Table S3 shows the comparison types for the independent patient groups (clinical) and 'PTSD-like' animal groups (preclinical).

***Table S3. Comparison types in clinical and preclinical data***

| | **Number of unique clinical PTSD groups (% of total 99 unique groups)** | **Number of unique preclinical PTSD groups (% of total 309 unique groups)** |
|---|---|---|
| **Experiment: Comparison Type** | | |
| non-exposed vs PTSD | 37 (37%) | 14 (5%) |
| non-exposed vs trauma-exposed | 2 (2%) | 288 (93%) |
| trauma-exposed vs PTSD | 60 (61%) | 7 (2%) |

Tables below (S4-4) show the characteristics of the independent PTSD groups that were included in the meta-analysis. Note, the *non-exposed vs trauma-exposed comparison (2 groups)* was not included in main analyses on clinical data (hypothesis testing and explorative analysis), since this experimental group does not contain actual PTSD patients. This comparison was only used in the clinical sensitivity analysis, as it is the most prevalent comparison type in preclinical data.

***Table S4. Characteristics clinical and preclinical data***

| | **Number of unique clinical PTSD groups (% of total 97 unique groups)** | **Number of unique preclinical PTSD groups (% of total 309 unique groups)** |
|---|---|---|
| **Experiment: Control Type** | | |
| Unexposed | 37 (38%) | 136 (44%) |
| Sham-exposed | 0 (0%) | 166 (54%) |
| Trauma-exposed | 60 (62%) | 7 (2%) |
| **Experiment: Time since trauma, (expressed in human years)** | | |

| | | |
|---|---|---|
| Mean (SD) | 15.73 (14.08) | 2.26 (3.04) |
| Range | 0.25 - 55.00 | 0.25 - 21.92 |
| Missing | 25 | 0 |
| **Experiment: Day/Night Phase** | | |
| active | 97 (100%) | 72 (23%) |
| inactive | 0 (0%) | 154 (50%) |
| unknown | 0 (0%) | 83 (27%) |
| **Subject: Sex** | | |
| Female | 9 (9%) | 15 (5%) |
| Male | 31 (32%) | 289 (94%) |
| Mixed | 56 (58%) | 3 (1%) |
| Missing | 1 | 2 |
| **Subject: Age** | | |
| (mature) adult | 27 (28%) | 36 (12%) |
| middle-aged | 54 (56%) | 0 (0%) |
| old adult | 16 (16%) | 0 (0%) |
| young adult | 0 (0%) | 273 (88%) |
| **Task: phase** | | |
| Learning | 44 (45%) | 56 (18%) |
| Memory | 50 (52%) | 226 (73%) |
| Extinction | 3 (3%) | 27 (9%) |
| **Task: Valence** | | |
| Neutral | 62 (64%) | 47 (15%) |
| Emotional | 10 (10%) | 0 (0%) |
| Fear | 24 (25%) | 121 (39%) |
| Trauma | 1 (1%) | 141 (46%) |
| **Task: Cue / Context** | | |
| Cue | 91 (94%) | 91 (29%) |
| Cue-Cue | 3 (3%) | 0 (0%) |
| Cue-Context | 2 (2%) | 2 (1%) |
| Context | 1 (1%) | 216 (70%) |
| **Task: information type** | | |
| autobiographical | 9 (9%) | 0 (0%) |
| olfactory | 0 (0%) | 1 (0%) |

| | | |
|---|---|---|
| safety | 3 (3%) | 4 (1%) |
| spatial | 1 (1%) | 38 (12%) |
| threat | 21 (22%) | 256 (83%) |
| verbal | 36 (37%) | 0 (0%) |
| verbal-visuospatial | 3 (3%) | 0 (0%) |
| visual | 18 (19%) | 10 (3%) |
| visual-verbal | 6 (6%) | 0 (0%) |

***Table S5. Characteristics clinical data***

| | **Number of unique clinical PTSD groups (% of total 97 unique groups)** |
|---|---|
| **Subject: Population** | |
| Civilian | 50 (52%) |
| Veterans | 47 (48%) |
| **Subject: Sample** | |
| Mixed | 9 (9%) |
| Military | 2 (2%) |
| Veteran | 39 (40%) |
| Military+Veteran | 1 (1%) |
| Civilian | 46 (47%) |
| **Subject: Trauma Sequelae** | |
| Accident | 1 (1%) |
| Assault | 5 (5%) |
| Childhood + Adulthood | 2 (2%) |
| Deployment | 44 (45%) |
| Deployment + Assault | 1 (1%) |
| Disaster | 7 (7%) |
| Mixed | 24 (25%) |
| Refugee | 3 (3%) |
| Unstated | 8 (8%) |
| WarRelated | 2 (2%) |
| **Task: Measure** | |
| non-physiology | 73 (75%) |

| | |
|---|---|
| physiology | 24 (25%) |

##### *Table S6. Characteristics preclinical data*

| | Number of unique preclinical PTSD groups (% of total 309 unique groups) |
|---|---|
| **Subject: Population** | |
| Albino Swiss (CD1) | 9 (3%) |
| Balb/c | 1 (0%) |
| C57BL/6 | 37 (12%) |
| Charles Foster | 2 (1%) |
| DBA/2 J | 2 (1%) |
| Fischer 344 | 1 (0%) |
| ICR | 1 (0%) |
| KunMing | 1 (0%) |
| Lewis | 4 (1%) |
| Long-Evans | 11 (4%) |
| OF1 | 2 (1%) |
| Sprague-Dawley | 186 (60%) |
| Swiss | 1 (0%) |
| Wistar | 51 (17%) |
| **Subject: Sample** | |
| Rat | 255 (83%) |
| Mice | 54 (17%) |
| **Subject: Trauma Sequelae** | |
| Footshock | 110 (36%) |
| Mixed | 20 (6%) |
| Predator Scent Stress | 50 (16%) |
| Restrained Stress | 16 (5%) |
| Social Defeat | 2 (1%) |
| Single Prolonged Stress | 106 (34%) |
| Under Water Trauma | 5 (2%) |
| **Task: Measure** | |
| exploration | 107 (35%) |

| | |
|---|---|
| freezing | 199 (64%) |
| manipulation | 3 (1%) |

#### B2. Study Quality: Risk of bias

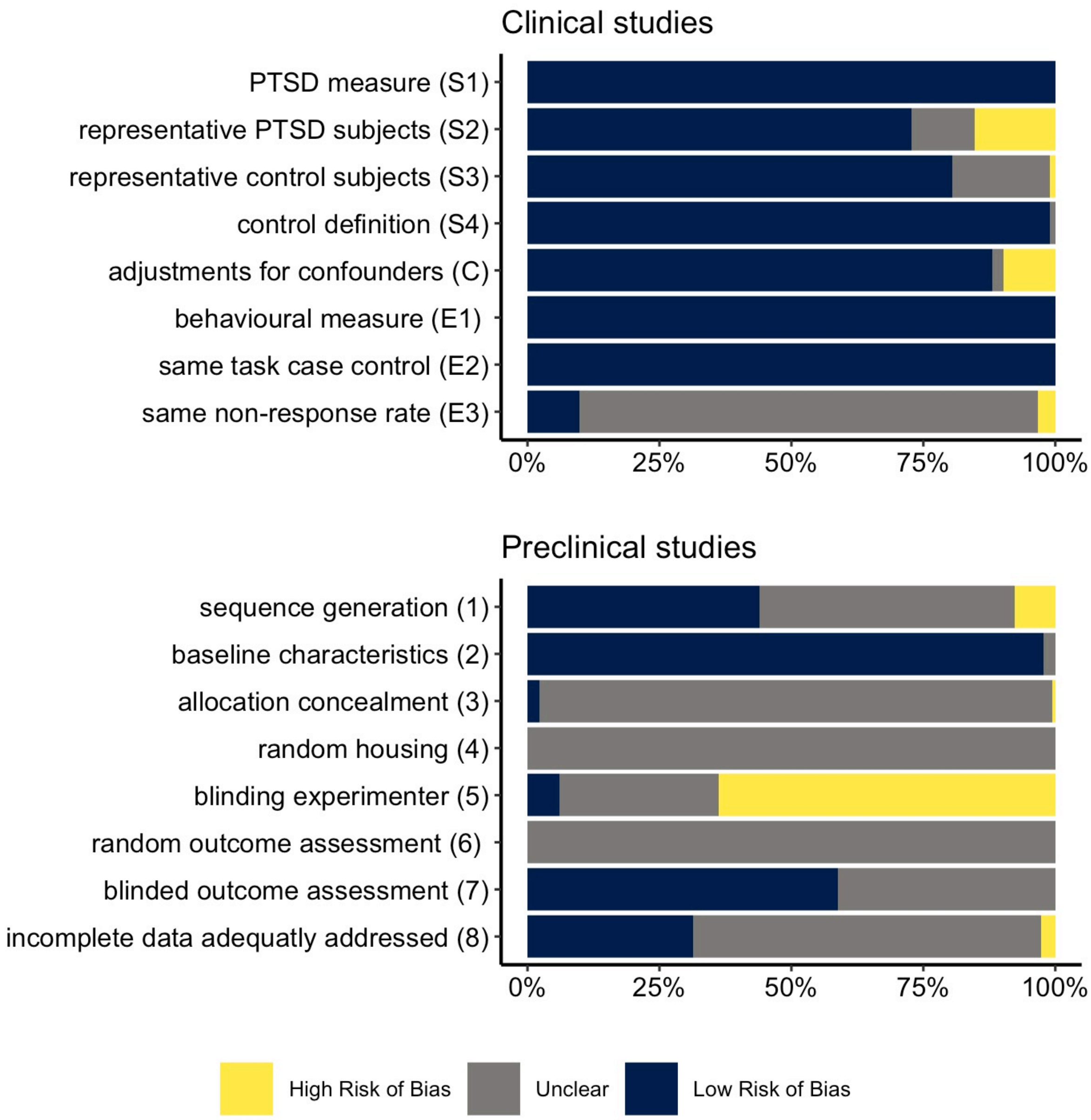

Figure S1. Risk of Bias.

The risk of bias in clinical studies was assessed on the *Newcastle-Ottawa case-control Scale* (3) items (top). The items of *SYRCLE's risk of bias tool* (4) were used to asses risk of bias in preclinical studies (bottom). Only studies that were included in the meta-analysis are displayed in these graphs.

#### B3. Results random-effects meta-regression

Table S7. Results random-effects meta-regression in clinical data

| Valence x phase | Hedge's G | standard error | lower bound 95%CI | upper bound 95%CI | Z | P uncorrected | P Bonferroni corrected |
|---|---|---|---|---|---|---|---|
| **neutral Learning** | -0,667 | 0,092 | -0,847 | -0,488 | -7,287 | <.001 | <.001 |
| **neutral Memory** | -0,544 | 0,087 | -0,715 | -0,373 | -6,239 | <.001 | <.001 |
| **fear Learning** | -0,200 | 0,155 | -0,503 | 0,103 | -1,295 | 0.196 | 1 |
| **emotional Memory** | -0,655 | 0,113 | -0,876 | -0,434 | -5,801 | <.001 | <.001 |
| **fear Extinction** | -0,804 | 0,158 | -1,114 | -0,494 | -5,086 | <.001 | <.001 |
| | **Difference in Hedge's G** | | | | | | |
| **Learning: neutral vs emotional** | 0,467 | 0,180 | 0,115 | 0,819 | 2,600 | 0.009 | 0.065 |
| **Memory: neutral vs emotional** | -0,111 | 0,086 | -0,28 | 0,058 | -1,291 | 0.197 | 1 |

Table S8. Results random-effects meta-regression in preclinical data

| Valence x phase | Hedge's G | standard error | lower bound 95%CI | upper bound 95%CI | Z | P uncorrected | P Bonferroni corrected |
|---|---|---|---|---|---|---|---|
| **neutral Learning** | -1,304 | 0,155 | -1,608 | -0,999 | -8,387 | <.001 | <.001 |
| **neutral Memory** | -1,291 | 0,152 | -1,588 | -0,994 | -8,518 | <.001 | <.001 |
| **fear Learning** | 0,435 | 0,146 | 0,15 | 0,72 | 2,987 | 0.003 | 0.034 |
| **fear Memory** | 0,812 | 0,136 | 0,545 | 1,079 | 5,962 | <.001 | <.001 |
| **fear Extinction** | -0,741 | 0,137 | -1,009 | -0,472 | -5,41 | <.001 | <.001 |
| **trauma Memory** | 1,877 | 0,138 | 1,606 | 2,148 | 13,563 | <.001 | <.001 |
| **trauma Extinction** | -2,19 | 0,23 | -2,64 | -1,739 | -9,532 | <.001 | <.001 |
| | **Difference in Hedge's G** | | | | | | |
| **Learning: neutral vs fear** | 1,738 | 0,172 | 1,402 | 2,075 | 10,12 | <.001 | <.001 |
| **Memory: neutral vs fear** | 2,103 | 0,159 | 1,792 | 2,414 | 13,261 | <.001 | <.001 |
| **Memory: neutral vs trauma** | 3,168 | 0,172 | 2,83 | 3,506 | 18,371 | <.001 | <.001 |
| **Memory: fear vs trauma** | 1,065 | 0,169 | 0,733 | 1,397 | 6,293 | <.001 | <.001 |
| **Extinction: fear vs trauma** | -1,449 | 0,253 | -1,944 | -0,954 | -5,731 | <.001 | <.001 |

#### B4. Robustness of the effects in random-effects meta-regression

##### *B4.1. Clinical data*

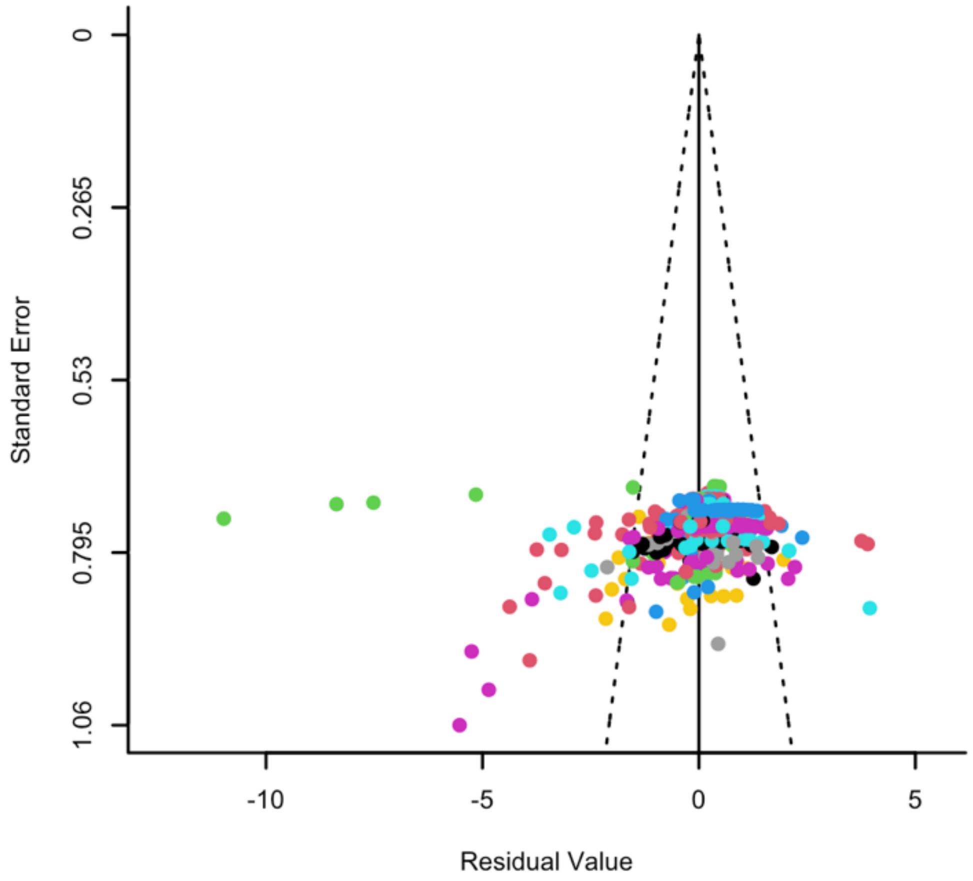

Figure S2. Funnel plot clinical data.

Egger's regression for funnel plot asymmetry: z = -4.798, p < .001.

Table S9. Rosenthal's fail-safe N for clinical data

| valence | phase | Fail-save N | P |
|---|---|---|---|
| Fear | Extinction | 10303 | <.001 |
| Fear | Learning | 1634 | <.001 |
| Neutral | Learning | 7914 | <.001 |
| Emotional | Memory | 4467 | <.001 |
| Neutral | Memory | 52014 | <.001 |

##### *B4.2. Preclinical data*

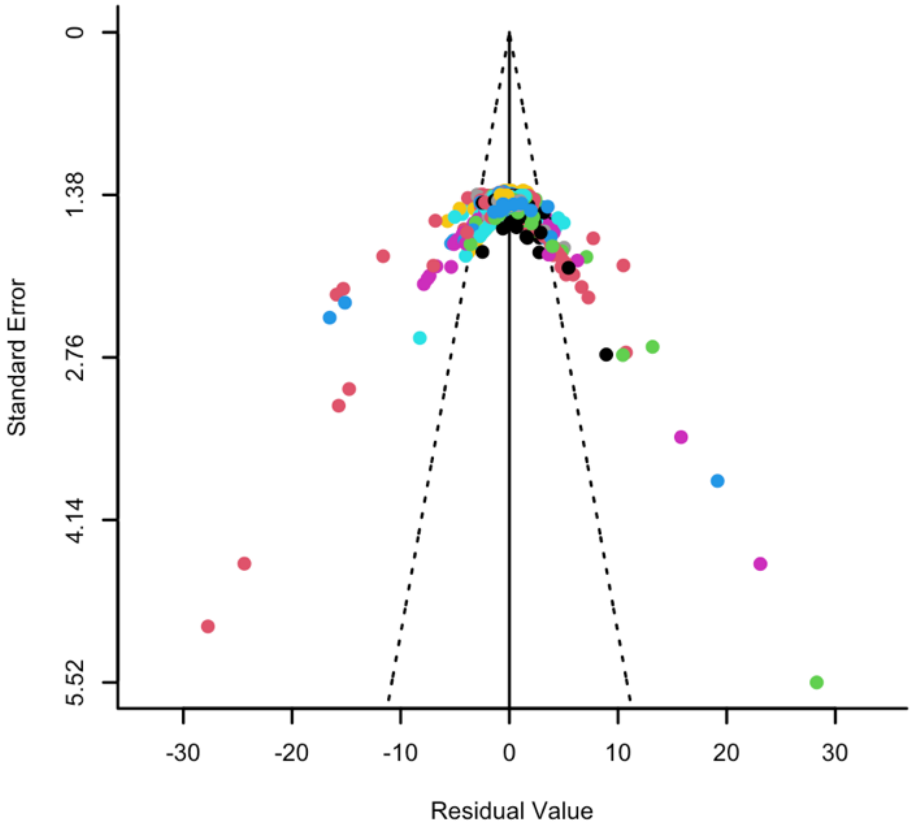

Figure S3. Funnel plot clinical data.

Egger's regression for funnel plot asymmetry: z = -1.326, p = 0.185

Table S10. Rosenthal's fail-safe N for preclinical data

| valence | phase | Fail-save N | P |
|---|---|---|---|
| Fear | Extinction | 84555 | <.001 |
| Trauma | Extinction | 10702 | <.001 |
| Fear | Learning | 867 | <.001 |
| Neutral | Learning | 35308 | <.001 |
| Fear | Memory | 27443 | <.001 |
| Neutral | Memory | 21941 | <.001 |
| Trauma | Memory | 265760 | <.001 |

### B5. Sensitivity analysis random-effects meta-regression

#### *B5.1. Clinical data*

##### Analysis without potential outliers and influential cases

17 comparisons were identified as potential outliers and influential cases (5), from 8 different clinical studies. Results of the random effects meta-regression without these cases are shown in Table S11.

**Table S11. Results meta-regression without potential outliers and influential cases in clinical data**

| Valence x phase | Hedge's G | standard error | lower bound 95%CI | upper bound 95%CI | Z | P |
|---|---|---|---|---|---|---|
| **fear Extinction** | -0,407 | 0,118 | -0,638 | -0,175 | -3,445 | 0.001 |
| **fear Learning** | -0,099 | 0,113 | -0,32 | 0,122 | -0,877 | 0.381 |
| **neutral Learning** | -0,664 | 0,069 | -0,8 | -0,528 | -9,584 | <.001 |
| **emotional Memory** | -0,52 | 0,095 | -0,706 | -0,335 | -5,494 | <.001 |
| **neutral Memory** | -0,549 | 0,064 | -0,674 | -0,425 | -8,638 | <.001 |

##### Analyses per comparison type

Clinical data was segregated per comparison type: non-trauma-exposed controls vs PTSD group (Table S12-S13); *non-trauma-exposed* controls *vs trauma-exposed* controls (Table S12-S15); and *trauma-exposed* controls *vs PTSD* group (Table S16-S17). The main random-effects meta-regression was subsequently repeated on the resulting three subsets.

**Table S12. Summary available data on comparison A in clinical data: non-trauma-exposed controls vs PTSD patients**

| phase | valence | papers | comparisons |
|---|---|---|---|
| Extinction | Fear | 6 | 41 |
| Learning | Fear | 12 | 43 |
| Learning | Neutral | 14 | 32 |
| Memory | Emotional | 7 | 21 |
| Memory | Neutral | 26 | 99 |

| | | | |
|---|---|---|---|
| Memory | Trauma | 1 | 1 |

Table S13. Results random effects meta-regression for comparison A in clinical data: non-trauma-exposed controls vs PTSD patients

| Valence x phase | Hedge's G | standard error | lower bound 95%CI | upper bound 95%CI | Z | P |
|---|---|---|---|---|---|---|
| **fear Extinction** | -0,511 | 0,226 | -0,954 | -0,069 | -2,266 | 0.023 |
| **fear Learning** | -0,387 | 0,219 | -0,817 | 0,042 | -1,767 | 0.077 |
| **neutral Learning** | -0,875 | 0,157 | -1,183 | -0,568 | -5,575 | <.001 |
| **emotional Memory** | -0,601 | 0,17 | -0,935 | -0,268 | -3,536 | <.001 |
| **neutral Memory** | -0,6 | 0,146 | -0,886 | -0,314 | -4,111 | <.001 |
| **trauma Memory** | 2,66 | 0,593 | 1,498 | 3,823 | 4,485 | <.001 |

Table S14. Summary available data on comparison B in clinical data: non-trauma-exposed controls vs trauma-exposed controls

| phase | valence | papers | comparisons |
|---|---|---|---|
| Learning | Neutral | 1 | 1 |
| Memory | Neutral | 2 | 4 |

Table S15. Results random effects meta-regression for comparison B in clinical data: non-trauma-exposed controls vs trauma-exposed controls

| Valence x phase | Hedge's G | standard error | lower bound 95%CI | upper bound 95%CI | Z | P |
|---|---|---|---|---|---|---|
| **Neutral Learning** | -0,126 | 0,438 | -0,985 | 0,733 | -0,288 | 0,773 |
| **Neutral Memory** | -0,275 | 0,136 | -0,541 | -0,008 | -2,018 | 0,044 |

Note, the available data on comparison B only includes one level of valence (neutral), therefore the variable valence is dropped from the model.

Table S16. Summary available data on comparison C in clinical data: trauma-exposed controls vs PTSD patients

| phase | valence | papers | comparisons |
|---|---|---|---|
| Extinction | Fear | 10 | 40 |
| Learning | Fear | 13 | 39 |
| Learning | Neutral | 29 | 63 |
| Learning | Trauma | 1 | 1 |
| Memory | Emotional | 10 | 28 |
| Memory | Fear | 1 | 2 |
| Memory | Neutral | 44 | 141 |
| Memory | Trauma | 2 | 2 |

Table S17. Results random effects meta-regression for comparison C in clinical data: trauma-exposed controls vs PTSD patients

| Valence x phase | Hedge's G | standard error | lower bound 95%CI | upper bound 95%CI | Z | P |
|---|---|---|---|---|---|---|
| **fear Extinction** | -1,022 | 0,188 | -1,389 | -0,654 | -5,446 | <.001 |
| **fear Learning** | 0,098 | 0,184 | -0,264 | 0,459 | 0,53 | 0.596 |
| **neutral Learning** | -0,545 | 0,104 | -0,748 | -0,341 | -5,234 | <.001 |
| **trauma Learning** | -1,161 | 0,298 | -1,745 | -0,577 | -3,895 | <.001 |
| **emotional Memory** | -0,854 | 0,159 | -1,167 | -0,541 | -5,354 | <.001 |
| **fear Memory** | 0,561 | 0,38 | -0,185 | 1,306 | 1,474 | 0.141 |
| **neutral Memory** | -0,476 | 0,099 | -0,67 | -0,281 | -4,8 | <.001 |
| **trauma Memory** | -0,101 | 0,285 | -0,66 | 0,457 | -0,355 | 0.722 |

##### *B5.2. Preclinical data*

#### Analysis without potential outliers and influential cases

15 comparisons were identified as potential outliers and influential cases (5), from 8 different preclinical studies. Results of the random effects meta-regression without these cases are in Table S18.

Table S18. Results meta-regression without potential outliers and influential cases in preclinical data

| Valence x phase | Hedge's G | standard error | lower bound 95%CI | upper bound 95%CI | Z | P |
|---|---|---|---|---|---|---|
| **fear Extinction** | -0,743 | 0,119 | -0,976 | -0,511 | -6,261 | <.001 |
| **trauma Extinction** | -2,136 | 0,214 | -2,555 | -1,718 | -10,004 | <.001 |
| **fear Learning** | 0,428 | 0,129 | 0,176 | 0,68 | 3,333 | 0.001 |
| **neutral Learning** | -1,251 | 0,138 | -1,522 | -0,979 | -9,035 | <.001 |
| **fear Memory** | 0,816 | 0,118 | 0,584 | 1,048 | 6,888 | <.001 |
| **neutral Memory** | -1,288 | 0,134 | -1,552 | -1,025 | -9,589 | <.001 |
| **trauma Memory** | 1,88 | 0,119 | 1,646 | 2,113 | 15,779 | <.001 |

#### Analyses per comparison type

Preclinical data was segregated per comparison type: *non-trauma-exposed* controls *vs trauma-exposed* controls (Table S19-S20); non-trauma-exposed controls vs PTSD group (Table S21-S22); and *trauma-exposed* controls *vs PTSD* group (Table S23-S24). The main random-effects meta-regression was subsequently repeated on the resulting three subsets.

Table S19. Summary available data on comparison D in preclinical data: non-trauma-exposed controls vs trauma-exposed controls

| phase | valence | papers | comparisons |
|---|---|---|---|
| Extinction | Fear | 44 | 242 |
| Extinction | Trauma | 7 | 68 |
| Learning | Fear | 30 | 79 |

| | | | |
|---|---|---|---|
| Learning | Neutral | 27 | 120 |
| Memory | Fear | 65 | 126 |
| Memory | Neutral | 45 | 94 |
| Memory | Trauma | 67 | 234 |

Note, in most preclinical studies, PTSD is modeled as 'trauma-exposed'.

Table S20. Results random effects meta-regression for comparison D in preclinical data: non-trauma-exposed controls vs trauma-exposed controls

| Valence x phase | Hedge's G | standard error | lower bound 95%CI | upper bound 95%CI | Z | P |
|---|---|---|---|---|---|---|
| **fear Extinction** | -0,718 | 0,143 | -0,999 | -0,438 | -5,019 | <.001 |
| **trauma Extinction** | -2,382 | 0,245 | -2,861 | -1,903 | -9,742 | <.001 |
| **fear Learning** | 0,442 | 0,152 | 0,144 | 0,74 | 2,909 | 0.004 |
| **neutral Learning** | -1,252 | 0,161 | -1,568 | -0,937 | -7,776 | <.001 |
| **fear Memory** | 0,828 | 0,142 | 0,549 | 1,107 | 5,812 | <.001 |
| **neutral Memory** | -1,298 | 0,156 | -1,604 | -0,992 | -8,317 | <.001 |
| **trauma Memory** | 1,9 | 0,148 | 1,609 | 2,191 | 12,804 | <.001 |

Table S21. Summary available data on comparison E in preclinical data: non-trauma-exposed controls vs animals with PTSD-like behavior

| phase | valence | papers | comparisons |
|---|---|---|---|
| Extinction | Fear | 1 | 12 |
| Learning | Fear | 1 | 2 |
| Memory | Fear | 2 | 9 |
| Memory | Trauma | 9 | 24 |

Table S22. Results random effects meta-regression for comparison E in preclinical data: non-trauma-exposed controls vs animals with PTSD-like behavior

| Valence x phase | Hedge's G | standard error | lower bound 95%CI | upper bound 95%CI | Z | P |
|---|---|---|---|---|---|---|
| **fear Extinction** | -1,323 | 0,821 | -2,931 | 0,286 | -1,612 | 0.107 |
| **fear Learning** | 0,472 | 0,889 | -1,271 | 2,216 | 0,531 | 0.595 |
| **fear Memory** | 0,451 | 0,811 | -1,139 | 2,042 | 0,556 | 0.578 |
| **trauma Memory** | 1,835 | 0,418 | 1,016 | 2,654 | 4,391 | <.001 |

Table S23. Summary available data on comparison F in preclinical data: trauma-exposed controls vs animals with PTSD-like behavior

| phase | valence | papers | comparisons |
|---|---|---|---|
| Extinction | Fear | 2 | 17 |
| Extinction | Trauma | 1 | 2 |
| Learning | Fear | 2 | 4 |
| Learning | Neutral | 1 | 16 |
| Memory | Fear | 4 | 9 |
| Memory | Neutral | 1 | 4 |
| Memory | Trauma | 10 | 27 |

Table S24. Results random effects meta-regression for comparison F in preclinical data: trauma-exposed controls vs animals with PTSD-like behavior

| Valence x phase | Hedge's G | standard error | lower bound 95%CI | upper bound 95%CI | Z | P |
|---|---|---|---|---|---|---|
| **fear Extinction** | -0,915 | 0,224 | -1,355 | -0,475 | -4,078 | <.001 |
| **trauma Extinction** | -0,959 | 0,469 | -1,877 | -0,04 | -2,045 | 0.041 |
| **fear Learning** | 0,236 | 0,3 | -0,352 | 0,823 | 0,787 | 0.432 |
| **neutral Learning** | -0,17 | 0,337 | -0,83 | 0,49 | -0,504 | 0.614 |

| | | | | | | |
|---|---|---|---|---|---|---|
| **fear Memory** | 0,355 | 0,224 | -0,085 | 0,794 | 1,581 | 0.114 |
| **neutral Memory** | 0,143 | 0,352 | -0,546 | 0,832 | 0,408 | 0.684 |
| **trauma Memory** | 0,96 | 0,152 | 0,661 | 1,259 | 6,301 | <.001 |

#### B6. Results MetaForest and MetaCART

##### B6.1. Convergence MetaForest

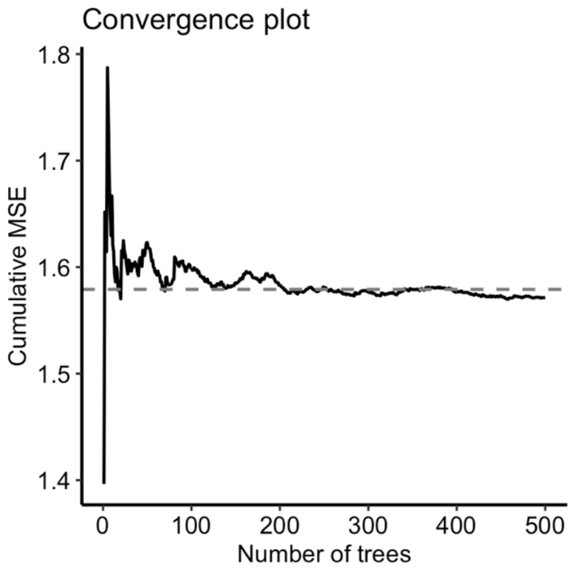

Figure S4. MetaForest convergence in clinical data

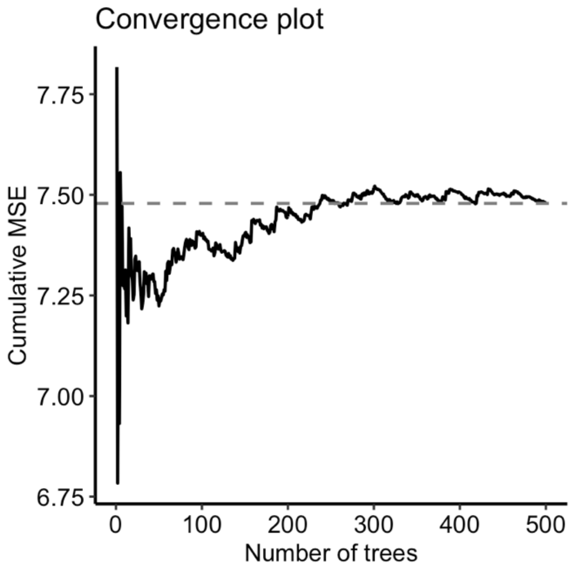

Figure S5. MetaForest convergence in preclinical data

#### *B6.2. METAFOREST AND META-CART PLOTS*

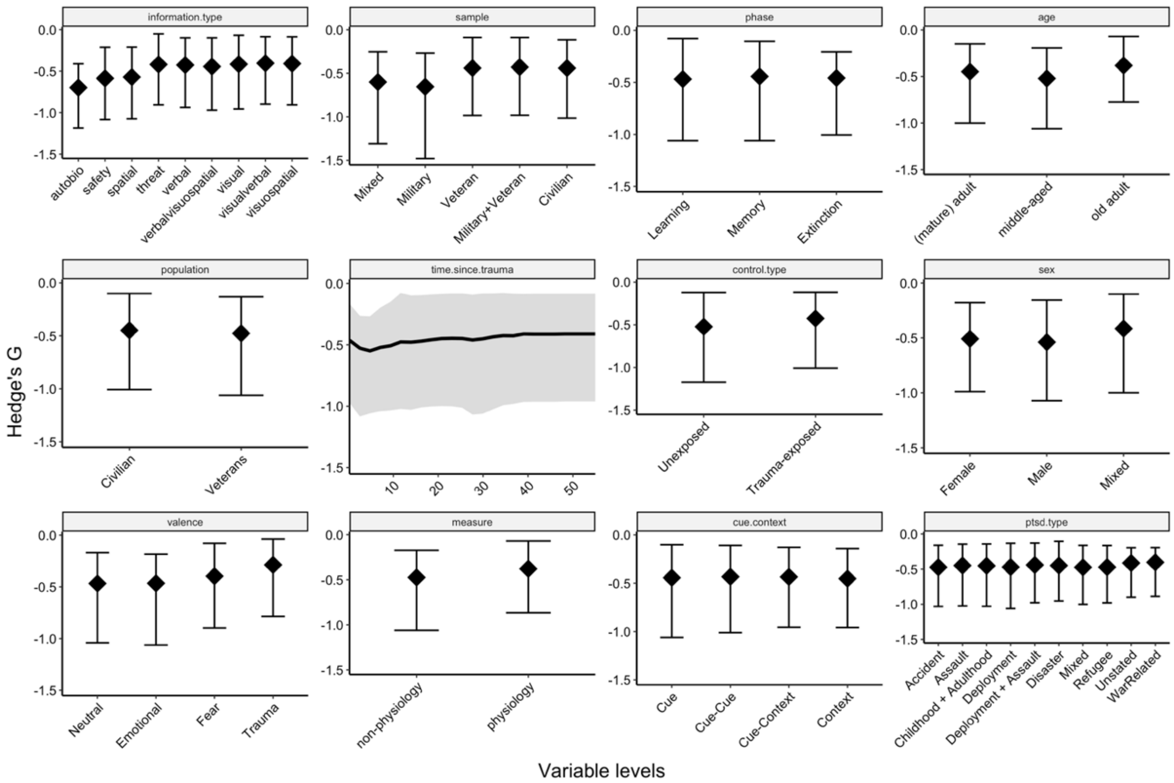

Figure S6. Partial dependence plots of clinical data

*Partial dependence plots showing the predicted relation (and 95% confidence interval) between Hedge's G and the variables in the random forest-based meta-analysis on clinical data.*

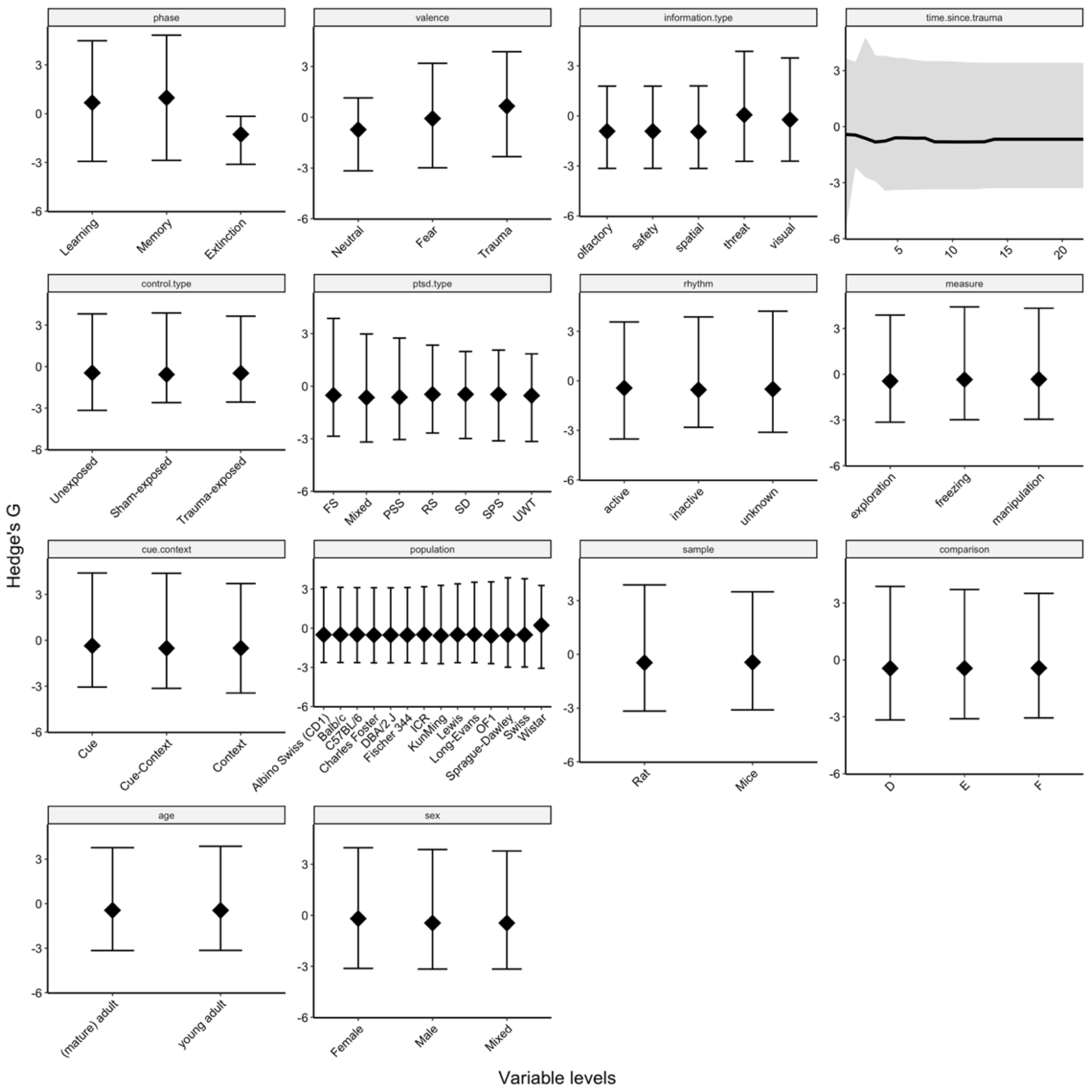

#### Figure S7. Partial dependence plots of preclinical data

*Partial dependence plots showing the predicted relation (and 95% confidence interval) between Hedge's G and the variables in the random forest-based meta-analysis on preclinical data.*

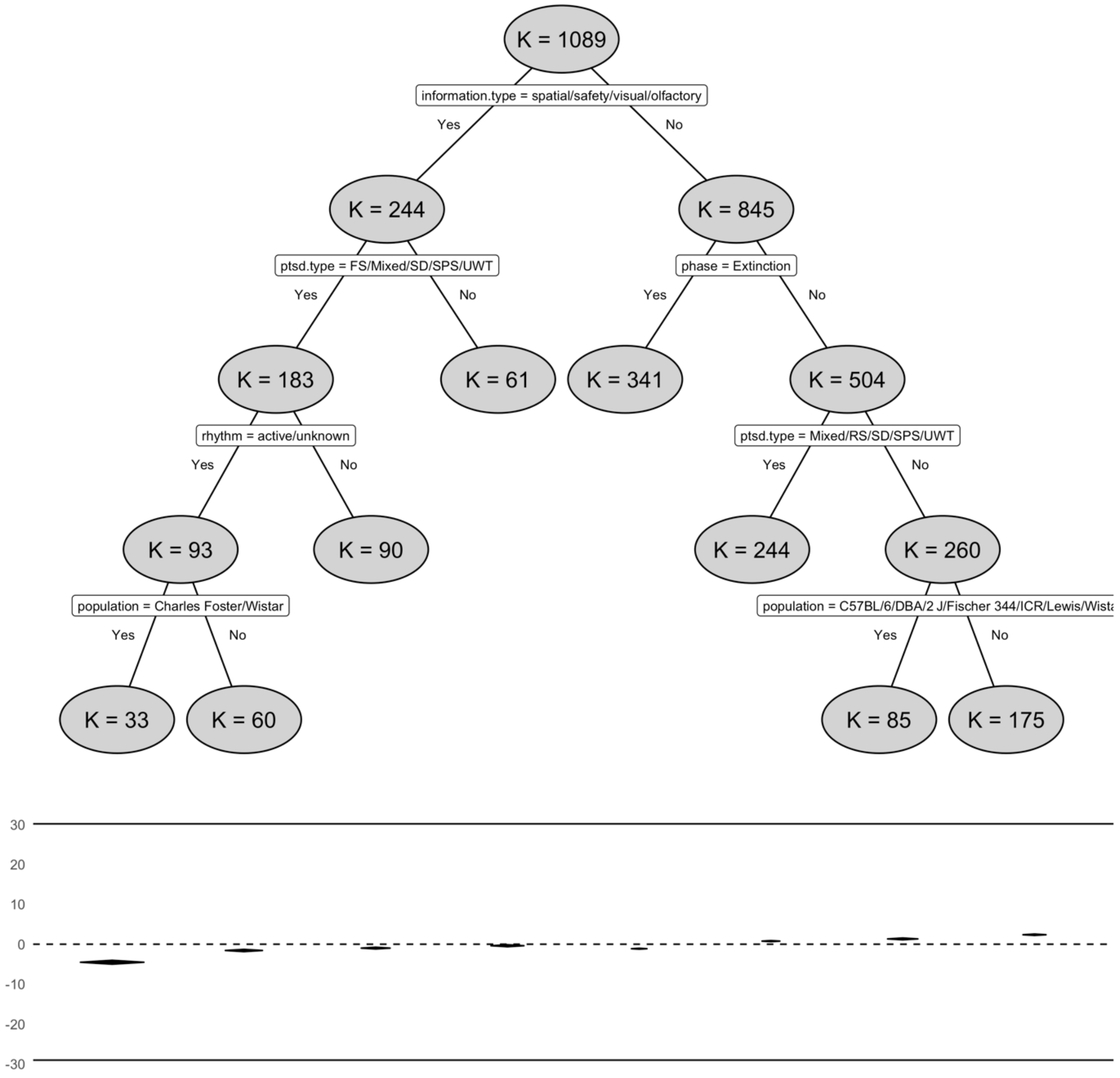

#### Figure S8. Partial dependence plots of Meta-CART interactions in preclinical data

Proposed interactions (upper) by Meta-CART and expected Hedge's G (lower).

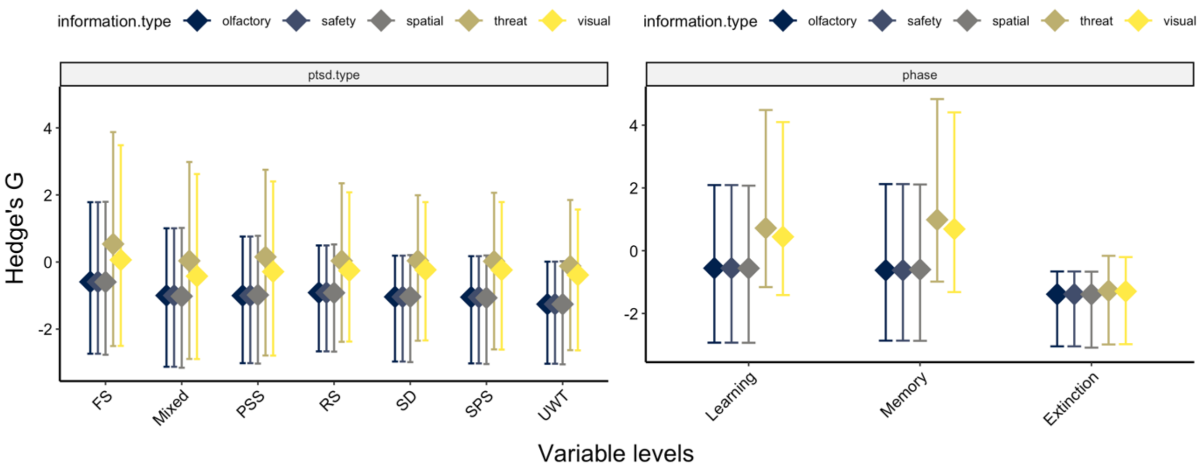

**Figure S9 Meta-CART preclinical in metaforest**

Partial dependence plots showing predicted Hedge's G (and 95% confidence interval) by upper Meta-CART interactions.
